# Race, Sex and Age Disparities in the Performance of ECG Deep Learning Models Predicting Heart Failure

**DOI:** 10.1101/2023.05.19.23290257

**Authors:** Dhamanpreet Kaur, John W. Hughes, Albert J. Rogers, Guson Kang, Sanjiv M. Narayan, Euan A. Ashley, Marco V. Perez

**Affiliations:** Stanford University; VA Palo Alto Health Care System

## Abstract

**Background:** Deep learning models may combat widening racial disparities in heart failure outcomes through early identification of individuals at high risk. However, demographic biases in the performance of these models have not been well studied.

**Methods:** This retrospective analysis used 12-lead ECGs taken between 2008 - 2018 from 290,252 patients referred for standard clinical indications to Stanford Hospital. The primary model was a convolutional neural network model trained to predict incident heart failure within 5 years. Biases were evaluated on the testing set (160,312 ECGs) using area under the receiver operating curve (AUC), stratified across the protected attributes of race, ethnicity, age, and sex.

**Results:** 50,956 incident cases of heart failure were observed within 5 years of ECG collection. The performance of the primary model declined with age. There were no significant differences observed between racial groups overall. However, the primary model performed significantly worse in Black patients aged 0 - 40 compared to all other racial groups in this age group, with differences most pronounced among young Black women. Disparities in model performance did not improve with integration of race, ethnicity, gender, and/or age into model architecture, by training separate models for each racial group, nor by providing the model with a dataset of equal racial representation. Using probability thresholds individualized for race, age, and gender offered substantial improvements in F1-scores.

**Conclusion:** The biases found in this study warrant caution against perpetuating disparities through the development of machine learning tools for the prognosis and management of heart failure. Customizing the application of these models by using probability thresholds individualized by race/ethnicity, age, and sex may offer an avenue to mitigate existing algorithmic disparities.

## Introduction

Heart failure (HF) remains one of the leading causes of death in the US, currently affecting 6.2 million adults^1^. The burden of the disease varies greatly by age, race, and gender^2^. Despite advances in medical care that have allowed incidence to stabilize or decline^3^, disparities in outcomes persist; in fact, the gap in age-adjusted death rates between Black patients and White patients has widened from 1999-2017, especially among younger patients^4^. HF may be underdiagnosed at higher rates in Black patients and women in the outpatient setting^5^. Earlier detection and closer monitoring of high-risk individuals may aid in reducing occurrence and improving prognosis of the disease to ultimately combat these disparities.

Several studies have used machine learning algorithms to identify cardiovascular conditions from electrocardiograms (ECGs)^6–8^. Deep neural networks can outperform cardiologists in recognizing several abnormalities from 12-lead ECG recordings, achieving F1- scores above 80% and specificity above 99%^9^. They can also achieve superior performance when compared to commercial rule-based methods, such as MUSE^10^. The application of artificial intelligence to ECG data has additionally been used to identify conditions that are not typically detected by the human eye. Deep learning models have been successfully developed to screen for asymptomatic left ventricular dysfunction (area under the receiver operator curve (AUC) of 0.93)^11^, atrial fibrillation (AUC of 0.87)^12^, aortic stenosis (AUC of 0.884)^13^ and anemia (AUC of 0.923)^14^, illustrating the broad potential of this non-invasive tool.

However, the performance and applicability of these models is highly contingent on the quality of the training data used, as well as the populations from which they are derived, and therefore may be prone to perpetuating implicit biases^15^. Several instances of this have been noted in the medical field: higher rates of underdiagnosis in chest radiographs among intersectional underserved populations^16^, significantly worse AUC values reported for dermatology AI algorithms when tested on diverse datasets^17^, lower risk scores for Black patients equally as ill as White patients^18^, and a 50% reduction in diagnostic accuracy of skin lesions among darker-skinned patients^19^. On the other hand, there have been cases of machine learning algorithms producing less biased results in comparison to other scoring methods^20^. Nonetheless, the presence of such biases remains largely understudied, particularly in the realm of machine learning applications for cardiovascular data.

In the case of ECG data specifically, disparities may be exacerbated by baseline differences due to age, race, and sex.^21, 22^ Several studies have particularly noted benign variations in ECG patterns among Black patients: ST segment flattening in young Black women^23^, greater ST-elevation thresholds in Black patients than White patients^24^, and biphasic T-waves in Black women^25^. The differences in ECG characteristics may also carry prognostic significance, showing inconsistent association with mortality, which holds implications for race-specific reference ranges and cardiovascular risk.^26^ Few studies have evaluated the disparities present in machine learning models applied to ECG data. Noseworthy et al. noted that in spite of racial differences in ECG data, a convolutional neural network trained on a homogenous population generalizes well to detecting low ejection fractions for several racial subgroups^27^. However, the effects of the intersectionality of age, sex, and race/ethnicity on model performance have not been well-studied.

This work aims to holistically investigate the existence of algorithmic biases as they pertain to age, race, ethnicity, and sex in a deep learning model trained to predict heart failure from ECG data and further explore how various modifications to the training and application of the model affect its performance.

## Methods

### Study Population and Data Sources

The 12-lead ECGs used in this retrospective analysis were derived from a total of 290,252 patients referred for standard clinical indications to Stanford University Medical Center. A total of 954,817 ECGs taken between March 2008 and May 2018 were extracted from the Phillips TraceMaster system. All ECGs were saved as 10 second signals from all 12 leads of the ECG, sampled at 500Hz. Band pass and wandering baseline filters were applied to the signals, normalized on a per-lead basis, and down sampled to 250Hz. During model evaluation, only the first ECG from each patient was considered.

Race, ethnicity, and sex were derived from self-report by the patients at the time of hospital enrollment. Follow-up heart failure from March of 2008 to February of 2022 was queried from STARR-OMOP^28^, a common data model for accessing electronic health records. Our primary outcome of interest was incident heart failure, defined as a first instance of heart failure within five years of the ECG. Heart failure was defined following prior work to include SNOMED code 84114007 (heart failure) and all descendants, excluding 82523003 (congestive rheumatic heart failure)^29^. Patients with prior heart failure were excluded. During training and testing, positive cases were defined as patients who developed heart failure within five years of the ECG; negative cases were defined as patients with no heart failure within five years of ECG, given at least five years of follow-up evidenced by measurement, admission, or mortality.

### Model development and training

A convolutional neural net was trained to predict occurrence of heart failure within 5 years of the ECG recording. Model development was performed using Python 3.9 and PyTorch 1.11, and models were trained on single Nvidia Titan Xp GPUs using Stanford’s Sherlock computing cluster. Hyperparameters were tuned by learning from the training set (88,084 ECGs) and evaluating on the validation set (28,882 ECGs). The parameters were set using a batch size of 128, a weight decay hyperparameter of 10^−4^, and the ADAM optimizer. The learning rate was initialized to 10^−3^ and reduced by a factor of 10 each time the validation loss plateaued for more than five epochs, limited at a lower bound of 10^−6^. After establishing hyperparameters, four more models were trained using cross-validation; the dataset was partitioned into five different subsets such that one of the five would be excluded from training and used as validation in each model. The outputs of these five models were averaged to generate predictions on the testing data (160,312 ECGs), which was then used for the analyses below.

### Statistical Analysis

A fair model would be expected to have equal predictive capacity across demographic divisions; the existence of group-based disparities in model performance constitutes bias. Age, race, ethnicity, and sex were chosen as the protected attributes across which to evaluate intersectional biases that may be present in model performance. Analyses were focused on the four largest racial and ethnic groups: Non-Hispanic White, Hispanic, Black, and Asian patients. Bias was defined as significant differences in area under the receiver operating curve (AUC) across demographic subgroups. Age was discretized into four groups of roughly equal proportions: 0-40 years, 40-60 years, 60-80 years, and >80 (80+) years. AUC was computed in a stratified manner for each demographic division. The receiver operating characteristic curves, AUC values and 95% confidence intervals were computed using the pROC R package^30^. Unless otherwise noted, AUC values were computed at a five-year time horizon, comparing all examples with an event of incident heart failure within five years against all examples with follow-up data after five years.

To assess differences in overdiagnosis versus underdiagnosis, calibration curves were graphed for each demographic subgroup using the Python Scikit-learn package^31^ as follows: the predicted probabilities were binned and plotted against the proportion of individuals in that bin that were observed to develop heart failure. The ideal model would show a 1:1 correlation (i.e., among individuals with predicted probabilities of 10%, 10% would be positive for observed occurrence of heart failure); a slope of less than one indicates overdiagnosis and a slope of greater than one indicates underdiagnosis.

The optimal threshold used in this model was based on F1-score using data from the entire population; precision, recall, and negative predictive value were computed within subgroups of race and gender. Moreover, the distribution of positive and negative ground truth labels for each subgroup was plotted against the probabilities assigned to the cases by the model.

### Reducing Model Biases

Three primary avenues in the design and application of the algorithm were investigated to reduce model biases: optimizing training data, modifying the architecture of the model, or adjusting application of the model.

In optimizing training data, the first approach involved training a separate model for each racial subgroup and each age subgroup. Four individual models were trained and tested on datasets consisting solely of Non-Hispanic White patients, Black patients, Asian patients, and Hispanic patients (e.g., the AUC value reported for Asian patients reflects performance of a model trained only on ECGs from Asian patients). Similarly, four individual models were trained and tested on datasets consisting solely of patients aged 0 - 40, 40 - 60, 60 - 80, and > 80. The second approach entailed providing the model with a dataset that has equal representation from each racial subgroup. Stratified sampling was applied to the full set of ECGs to take the same number of patients from each of the predominant four racial groups: using the size of the smallest group (Black patients) as the sampling quantity, this produced a test set of 27,176 ECGs.

In modifying the architecture of the model, the variables of race/ethnicity, age, and gender were incorporated through early and late fusion. In early fusion, the model is passed demographic data as an additional channel of input data. For example, a model trained on ECG and gender would be passed 13 channels of data, where the first 12 correspond to the 12-lead ECG and the 13^th^ is 0 where the patient is male and 1 if the patient is female. In late fusion, the ECG is first encoded as a low-dimensional latent vector to which demographic data is appended. This representation is then fed through fully connected layers to make final predictions. AUC values were compared across the demographic subgroups to validate the effectiveness of these approaches in reducing model bias.

In adjusting the application of the model, the probability threshold for classification of a case as a positive label was optimized separately for each demographic subgroup based on F1- score. For each demographic subgroup, the differences were computed between metrics (precision, recall, negative predictive value) that resulted from using the threshold value individualized for that group versus the overall optimal threshold value.

## Results

### Study Population

There were 290,252 patient ECGs derived from unique patients who were followed for an average of 6.8 years. The baseline characteristics stratified by incident heart failure are described in Table 1. The patients were on average 59.3 years old and comprised 49.7% women. The racial and ethnic composition consisted of Non-Hispanic White (56.5%), Asian (14.2%), Black (4.0%), Hispanic (12.3%), Native Hawaiian/Pacific Islander (1.2%), and American Indian or Alaskan Native (0.2%).

**Table 1.** Baseline characteristics (Hypertension defined as diastolic >80 or systolic >130; hyperlipidemia defined as LDL > 160)

| Demographic | Subgroup | All<br>(% of Total) | No Heart Failure<br>(% of Subgroup) | Heart Failure<br>(% of Subgroup) |
| --- | --- | --- | --- | --- |
| Age | 0 - 40 | 42433 (14.6%) | 38733 (91.3%) | 3700 (8.7%) |
|  | 40 - 60 | 95456 (32.9%) | 83924 (87.9%) | 11532 (12.1%) |
|  | 60 - 80 | 119537 (41.2%) | 95479 (79.9%) | 24058 (20.1%) |
|  | ≥ 80 | 32808 (11.3%) | 21143 (64.4%) | 11665 (35.6%) |
| Race | Asian | 41073 (14.2%) | 33890 (82.5%) | 7183 (17.5%) |
|  | Hispanic | 35806 (12.3%) | 29808 (83.2%) | 5998 (16.8%) |
|  | Non-Hispanic<br>White | 164081 (56.5%) | 134097 (81.7%) | 29984 (18.3%) |
|  | Black or African<br>American | 11471 (4.0%) | 8797 (76.7%) | 2674 (23.3%) |
|  | Native Hawaiian/<br>Pacific Islander | 3567 (1.2%) | 2655 (74.4%) | 912 (25.6%) |
|  | American Indian/<br>Alaskan Native | 578 (0.2%) | 503 (87.0%) | 75 (13.0%) |
| Gender | Male | 145892 (50.3%) | 117611 (80.6%) | 28281 (19.4%) |
|  | Female | 144354 (49.7%) | 121679 (84.3%) | 22675 (15.7%) |
| BMI | < 18.5 | 4852 (1.7%) | 3923 (80.9%) | 929 (19.1%) |
|  | 18.5 - 24.9 | 59042 (20.3%) | 47956 (81.2%) | 11086 (18.8%) |
|  | 25.0 - 29.9 | 60629 (20.9%) | 48180 (79.5%) | 12449 (20.5%) |
|  | ≥ 30.0 | 51481 (17.7%) | 39501 (76.7%) | 11980 (23.3%) |
| Hypertension | Positive Hx | 96485 (33.4%) | 77484 (80.0%) | 19361 (20.0%) |
| Diabetes | Positive Hx | 39018 (13.4%) | 27110 (69.5%) | 11908 (30.5%) |
| CAD or MI | Positive Hx | 38608 (13.3%) | 24913 (64.5%) | 13695 (35.5%) |
| Stroke | Positive Hx | 13083 (4.5%) | 9943 (76.0%) | 3140 (24.0%) |
| Hyperlipidemia | Positive Hx | 1698 (0.6%) | 1449 (85.3%) | 249 (14.7%) |
| Smoking | Positive Hx | 2012 (0.7%) | 1573 (78.2%) | 439 (21.8%) |

### Incidence of Heart Failure

There were 50,956 incident cases of heart failure within 5 years of ECG collection. The fraction of patients who developed heart failure within 5 years increased from 8.7% in the youngest age group of 0-40 year-olds to 35.6% in those over 80 years of age. The fraction of women (15.7%) developing heart failure within 5 years was lower than that of men (19.4%). The incidence of 5-year heart failure was higher among Black patients (23.3%) than White patients (18.3%), Hispanic patients (16.8%), or Asian patients (17.5%) (Table 1). Breakdowns of incidence rates by race/ethnicity, age, and gender can be found in Supplementary Table 1.

### Comparisons of Model Performance

The performance of the primary model declined significantly with age. The model performed significantly better in those 0 - 40 years old (AUC 0.80 [0.79 - 0.81]) compared to those who were >80 years old (AUC 0.66 [0.65 - 0.66]) (Figure 1a). The model performed slightly worse in men (AUC 0.77 [0.77 - 0.77]) compared to women (AUC 0.78 [0.78 - 0.79]) (Figure 1b). There were no significant differences observed in model performance between racial groups (Hispanic patients AUC 0.79 [0.79 - 0.80], Asian patients 0.78 [0.77 - 0.79], Non-Hispanic White patients 0.77 [0.77 - 0.78], Black patients 0.78 [0.77 - 0.79]) (Figure 1c).

**Figure 1.**
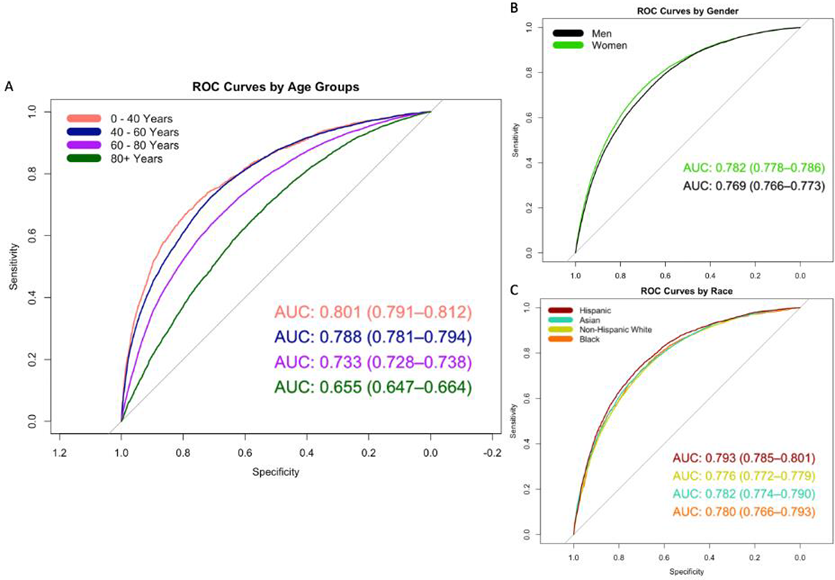
Receiver operator characteristic (ROC) curves and area under the curve (AUC) stratified by race, age, and gender

However, the trends in race- and gender-based disparities differed when broken down by age group. The primary model performed significantly worse in Black patients aged 0 - 40 years old (AUC 0.69 [95% CI 0.64 - 0.75]) compared to all other racial groups in the same age group (Non-Hispanic White AUC 0.80 [0.78 - 0.81]; Hispanic AUC 0.81 [0.79 - 0.83]; and Asian AUC 0.82 [0.79 - 0.85]) (Figure 2). The racial differences in AUC were much less pronounced among men, whereas the AUC for Black women (AUC 0.69 [0.62 - 0.77]) was substantially lower than women of all other racial groups aged 0 - 40 years (Supplementary Figure 1). Gender-based differences varied by age group (Supplementary Figure 2).

**Figure 2.**
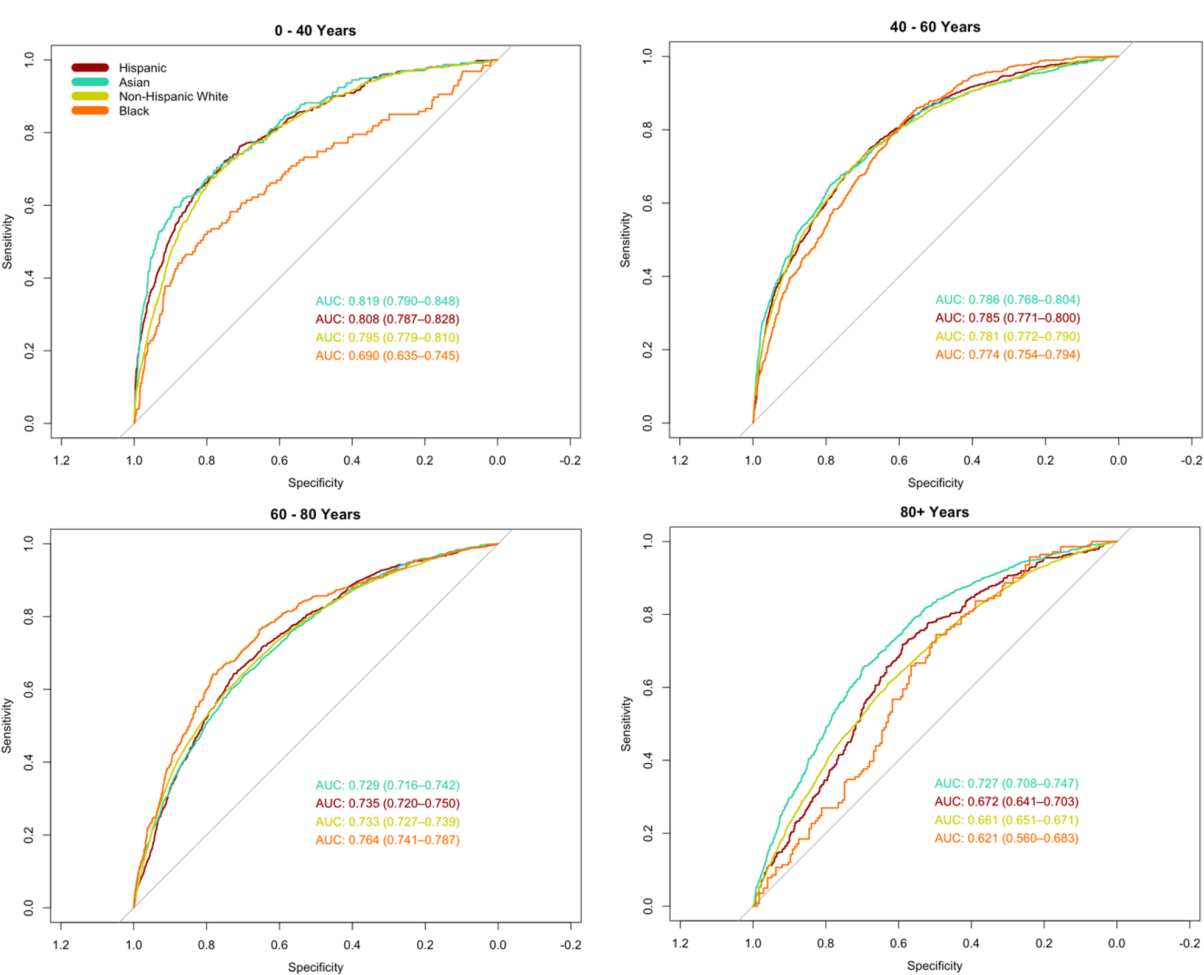
Receiver operator characteristic (ROC) curves and area under the curve (AUC) stratified by race across age groups

The distributions of predicted probabilities for positive and negative labels vary between race and age subgroups (Supplementary Figure 3). The overlap of predicted model probabilities between healthy patients and those who developed heart failure increased substantially with age across all racial groups. Among Black patients aged 0 - 40 years, a lower predicted probability is more likely to correlate with incident heart failure compared to other race and gender subgroups.

The calibration curves indicate that the model is best calibrated for Asian and Non-Hispanic White patients, as observed by the linear relationship between the predicted and observed probabilities. The observed fraction of cases with heart failure exceeds the probability predicted by the model among Black patients, indicating greater underdiagnosis in comparison to other racial groups, especially among Black women (Figure 3).

**Figure 3.**
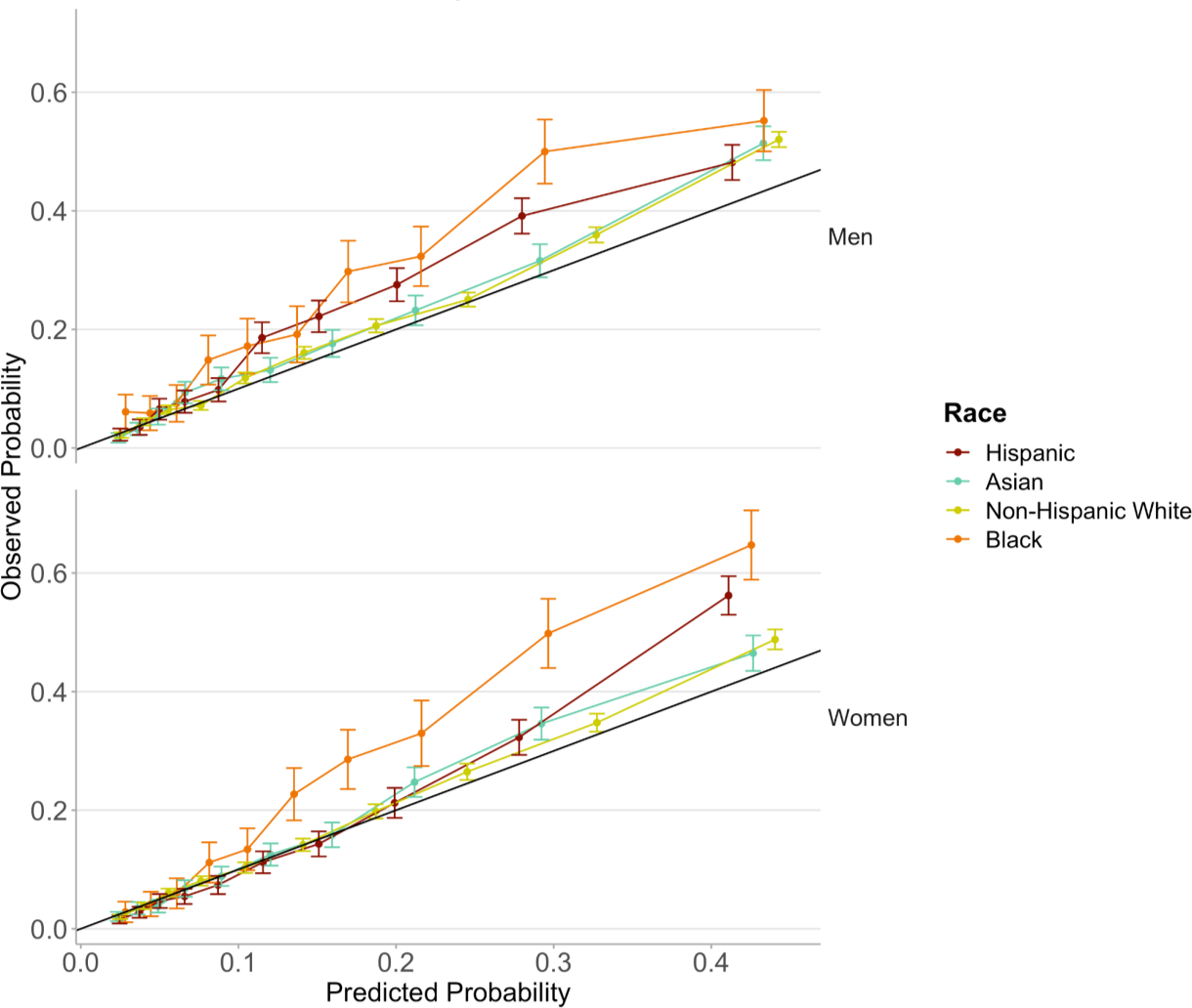
Calibration curves by race and gender

Figure 4 illustrates cases reflective of trends observed in model performance: an ECG from a Non-Hispanic White man who was assigned a high predicted probability and developed heart failure (true positive); an ECG from a Non-Hispanic White woman who was assigned a low predicted probability and did not develop heart failure (true negative); an ECG from a Black woman who was assigned a low predicted probability but developed heart failure (false negative); an ECG from an Asian man who was assigned a high predicted probability but did not develop heart failure (false positive); an ECG from a young Black woman with T-wave inversion who was assigned a high predicted probability but did not develop heart failure; and an ECG from a patient over 80 years old – showing typically low voltages and flat T-waves – who was assigned a high predicted probability but did not develop heart failure.

**Figure 4.**
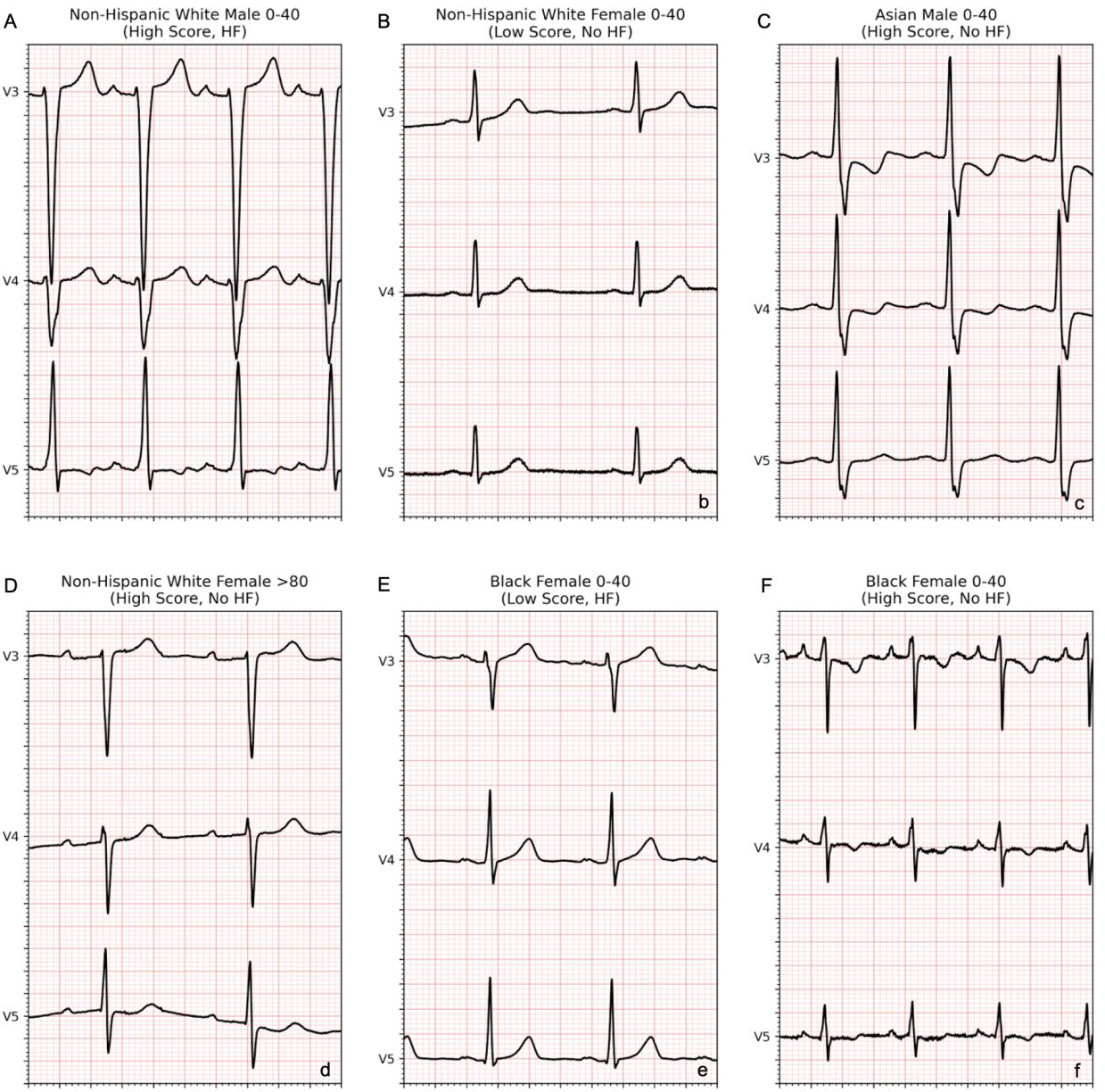
Sample ECGs showing a) Non-Hispanic White man with high predicted probability who developed HF, b) Non-Hispanic White woman with low predicted probability who did not develop HF, c) Asian man with high predicted probability who did not develop HF, d) Non-Hispanic White woman aged >80 years with high predicted probability who did not develop HF, e) Black woman aged 0-40 with low predicted probability who developed HF, and f) Black woman aged 0-40 with high predicted probability who did not develop HF

### Incorporating Race and Ethnicity in Model Building

The model was provided with different subsets of training data to determine if the disparities may have resulted from differences in racial and ethnic group sample sizes. Using a dataset with equal racial representation did not eliminate disparities between Black patients and patients of other racial groups in the 0 - 40 age group (Supplementary Figure 4). The AUC values did not improve from the primary model. Similarly, there was no improvement in performance amongst the different race and ethnic subgroups when compared using models that were trained on the same race and ethnicity as the test set (Supplementary Figure 5). Moreover, there was no improvement in age-related disparities when the models were trained and tested on data from separate age groups (Supplementary Figure 6).

The incorporation of race/ethnicity, age, gender, or the combination of those three demographic variables into training did not significantly improve performance in any subgroup of race and age. Nonetheless, there was a reduction in racial disparities among young patients from the combined effect of improving performance for Black patients and diminishing performance for other racial groups (Figure 5).

**Figure 5.**
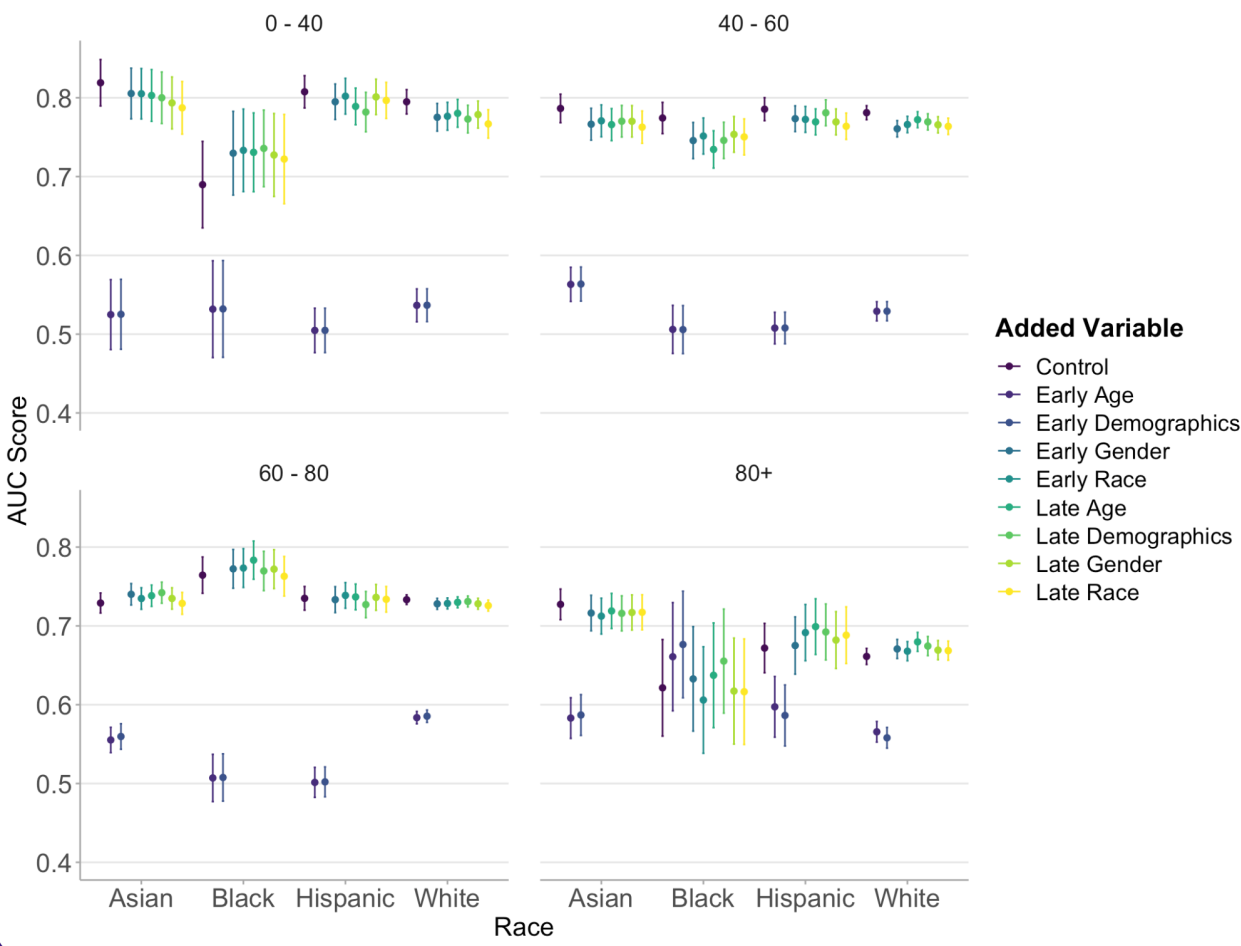
AUC values by race and age groups with integration of age, race, and gender variables at earlier and later points in model architecture

When factoring for F1-score, the optimal model probability threshold varied between subgroups of race/ethnicity, age, and gender (Supplementary Figure 7). Black men and women in the 0 - 40 and 40 - 60 year age groups had a lower optimal probability score threshold than all other groups. The optimal probability threshold for the entire population was 0.20; the highest individualized optimal threshold was 0.30 for Asian men aged 0 - 40 years and the lowest was 0.10 for Black men aged >80 years. All racial and gender groups aged 0 - 40 years showed increases in F1-scores when using individualized thresholds (Figure 6). Black women in this age group showed the greatest improvement using an individualized threshold of 0.15 with an 11%- point increase in F1-score – a 3% increase in PPV and a 21% increase in recall. Black men aged >80 years showed a 10% increase in F1-score using an individualized threshold of 0.10 – a 1%- point decrease in PPV and a 27%-point increase in recall (Supplementary Table 2).

**Figure 6.**
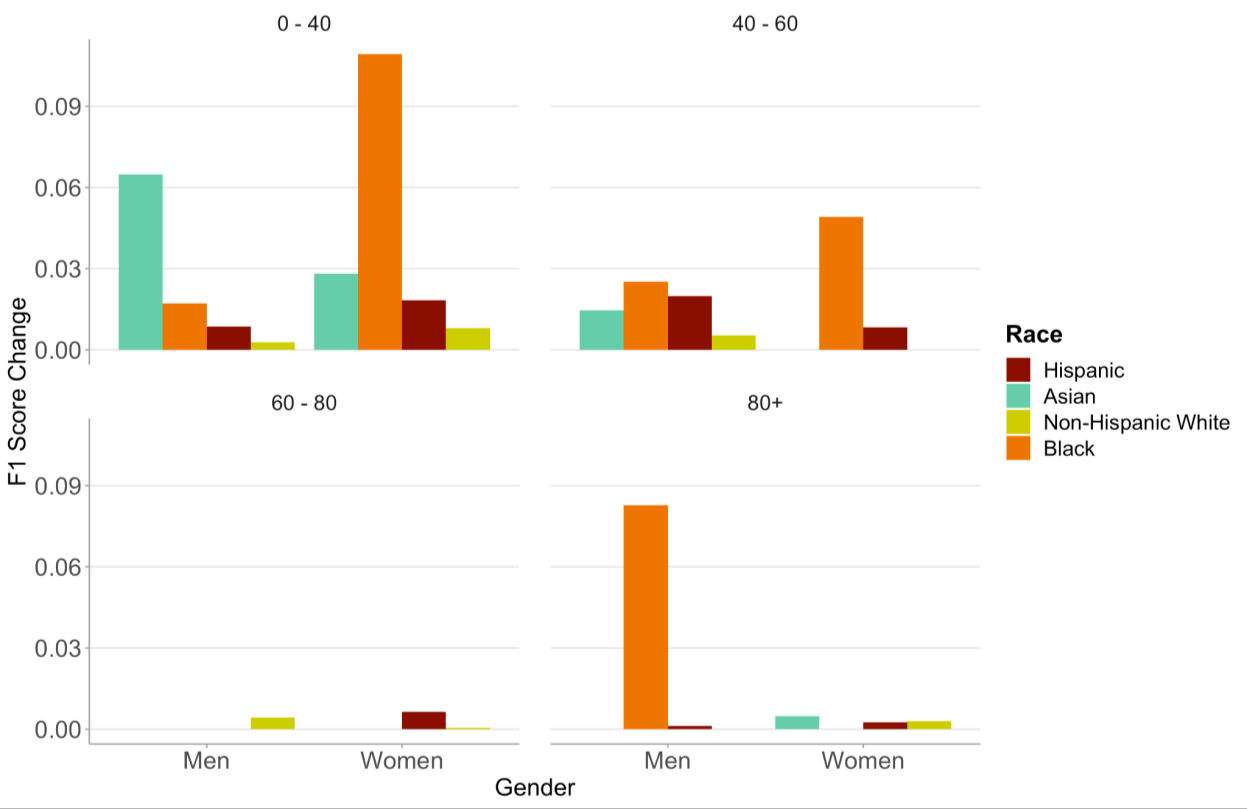
Changes in F1-Score when using probability thresholds individualized for race, age, and gender subgroups

## Discussion

The utility of deep learning models for EKG data arises from a reduction in the demand for cardiologist labor, comparable or increased accuracy of the automated output, and the ability to provide insights above and beyond those typically detected by the human eye. They provide clinicians with a powerful tool in advising treatment and predicting prognosis.

In this study, we assessed the algorithmic biases present in a model designed to detect 5- year occurrence of heart failure. We used a range of approaches previously proposed to assess algorithmic equity: predictive parity, predictive equality (false positive error rate balance), equal opportunity (false negative error rate balance), equalized odds, conditional use accuracy quality, and Well-calibration^32^. Some of the proposed standards are mathematically incompatible (i.e., fairness in one metric may preclude fairness in another)^33^, underscoring the need for a holistic evaluation and consideration of factors that influence utility of the model’s application.

The differences in AUC across demographic subgroups confirms the hypothesis that disparities exist in model performance, and further indicates that these disparities reflect the intersectionality of race/ethnicity, gender, and age in cardiovascular disease. Using the metric of AUC, we found that the model performed slightly better for women than men, and significantly worse among the older population and the Black population aged 0 - 40 than all other races. The small observed difference between men and women is notable given that there was a higher percentage of positive labels among men and consequently, less class imbalance in the data, which would be expected to enhance model performance^34^.

Upon further analyzing subgroups of race and gender, we found that the disparity observed among young Black patients stems primarily from the model’s diminished performance among Black women in comparison to women of other racial groups aged 0 - 40; significant differences do not exist between races among male patients or between Black men and Black women aged 0 - 40. There have been several cases of ECG patterns observed among healthy Black women that would typically be considered malignant in the White population^25^. This may be a contributing factor given the training data was predominantly comprised of White patients.

We found that model performance was worse in older patients compared to younger patients. Given that the composition of the training data was biased toward older patients and that older patients had a larger fraction developing heart failure, the most likely explanation for this trend is that there were more obvious ECG differences between healthy and diseased patients in the younger age groups.

Given the notable reduction in model performance pertaining to race and gender, we investigated multiple avenues of modifying the training and application of the model to improve the existing disparities. First and foremost, we acknowledge the widespread debate surrounding the use of race as a variable in algorithms for medical decision making.^35–37^ One study noted that race-specific models in the prediction of heart failure risk demonstrated superior performance to non-race-specific and traditional risk models.^38^ In this study, we do not seek to draw conclusions on whether race should be an input into prognosis of cardiovascular disease, but rather to investigate this as a method of improving predictions for minority groups. We did not find that the inclusion of race, gender, or age as variables in model training improved overall performance of the model. We did observe some reduction in racial disparities among the 0-40 age group, but this was due to the combined effects of improved performance in Black patients and diminished performance in the other racial groups. Models trained and tested on a dataset with equal racial representation or on datasets with individual race and age groups did not eliminate disparities in performance. However, it is difficult to discern the effects of the separation by race or age and stratified sampling from the decreased quantity of training data. Nevertheless, we believe that these findings warrant caution in using ML modeling as a tool for ECG interpretation and heart failure prognosis in older patients or in young, Black patients.

While AUC was a useful metric for holistically assessing model performance, we considered F1-score to evaluate the utility of the model in cases where a single binary cutoff is chosen for classification. The variance in thresholds at which F1-score peaks for different demographic subgroups suggests that individualized applications of the model may be optimal. The range in optimal threshold cutoffs varied from 0.10 in Black and Hispanic men aged >80 to 0.30 in Asian men aged 0-40. These threshold choices are supported by the distributions of positive case labels: in Black and Hispanic patients aged >80, positive cases follow a similar distribution to negative cases, so a lower threshold is necessary to capture more of those developing heart failure, whereas in young Asian men, the distribution of positive case labels is skewed more strongly toward higher probabilities. Moreover, the calibration curves showed overdiagnosis being most prominent among Black and Hispanic men, further concurring with a higher optimal threshold for these subgroups. In this study, we used F1-Score as the criteria for optimization, which gives equal weight to precision and recall. In doing so, we found a 21.0% increase in recall (the percentage of heart failure cases identified by the algorithm) for Black women. When applying such a model to the healthcare setting, we recognize that there may be greater value in recall than precision and recommend that the user assign an appropriate utility function for optimization. Thus, calibrating the algorithm for race, age, and gender after training may offer a promising avenue to reduce the disparities otherwise present in model performance.

### Limitations

The primary limitations of the study lie around the parameters of the outcome and cohort used to develop the model. We only considered patients with at least five years of follow-up data within the Stanford health system from the time of ECG collection. The exclusion of cases due to lack of follow-up may be biased by demographic group. However, one might expect that limiting the cohort to patients who sought longitudinal care within the Stanford system would create greater homogeneity in the data, thus rendering the findings of disparity in model performance even more striking. Furthermore, we assumed that any misclassification of cases would be non-differential based on demographic status. In the case that the misclassification is biased, we would expect higher rates of underdiagnosis for Black patients and women^5^, which only amplifies the finding of model underdiagnosis observed particularly for that group. Additionally, racial groups with limited sample sizes (Native Hawaiian/Pacific Islander and American Indian/Alaskan Native patients) were not included in the analyses so the results may not generalize to these populations.

## Conclusion

We analyzed demographic disparities on the basis of race, ethnicity, age, and gender present in a machine learning model trained to predict occurrence of 5-year heart failure from ECG data and found that performance suffered significantly for young Black women. We explored the mechanisms underlying these disparities through statistical covariates, calibration curves, and distributions of predicted probabilities. We investigated methods of improving these disparities – manipulation of the training data, modifications to model architecture, and optimization of threshold choice. This study serves to highlight the need to consider differential performance of machine learning models among demographic subgroups, and offers a framework for understanding, investigating, and improving the disparities that may otherwise be perpetuated by algorithms used for medical decision making.

## Data Availability

Individual patient data cannot be shared due to privacy concerns

## Acknowledgements

We do not have any acknowledgements to make.

## Supplemental Materials

**Supplementary Table 1.**
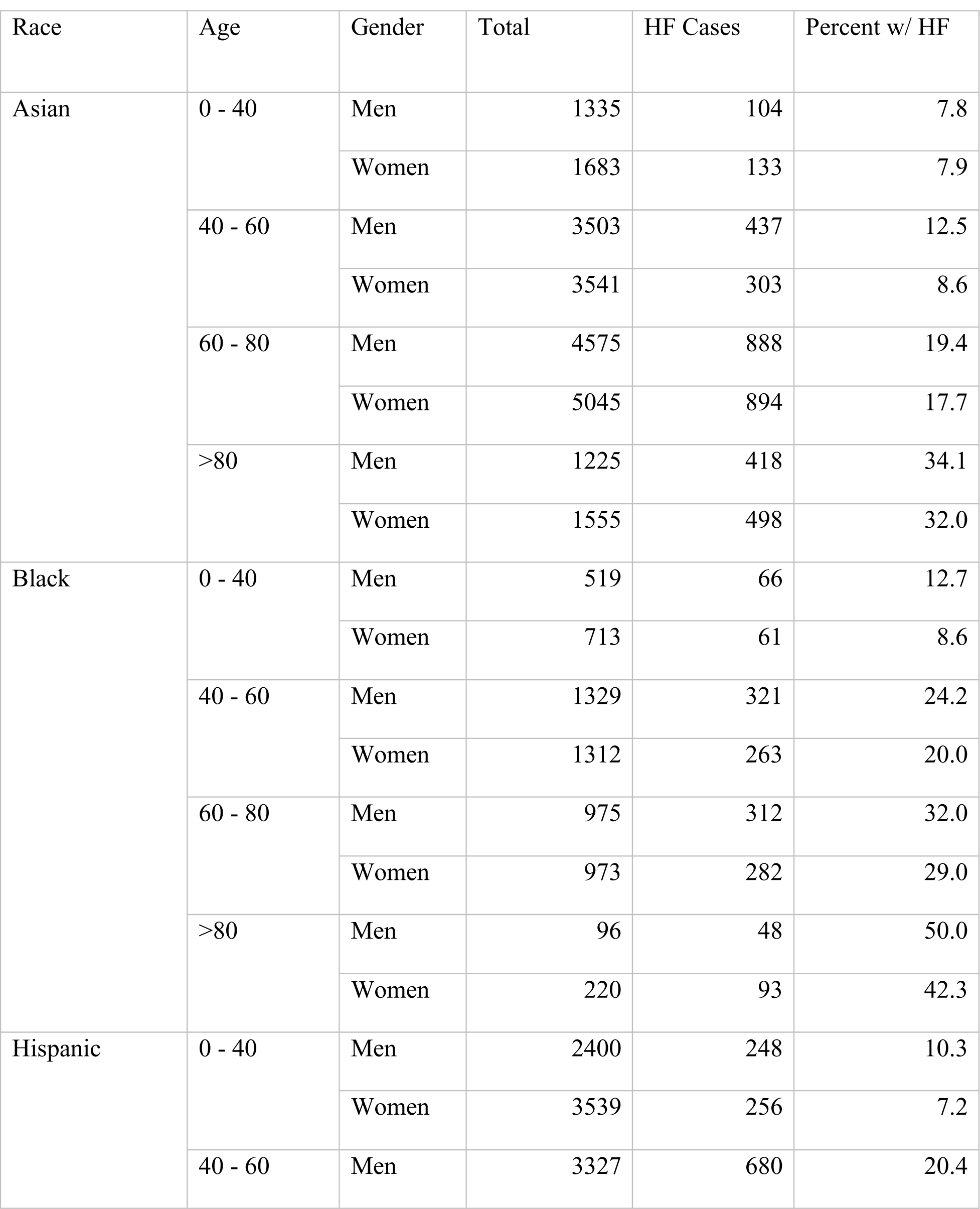
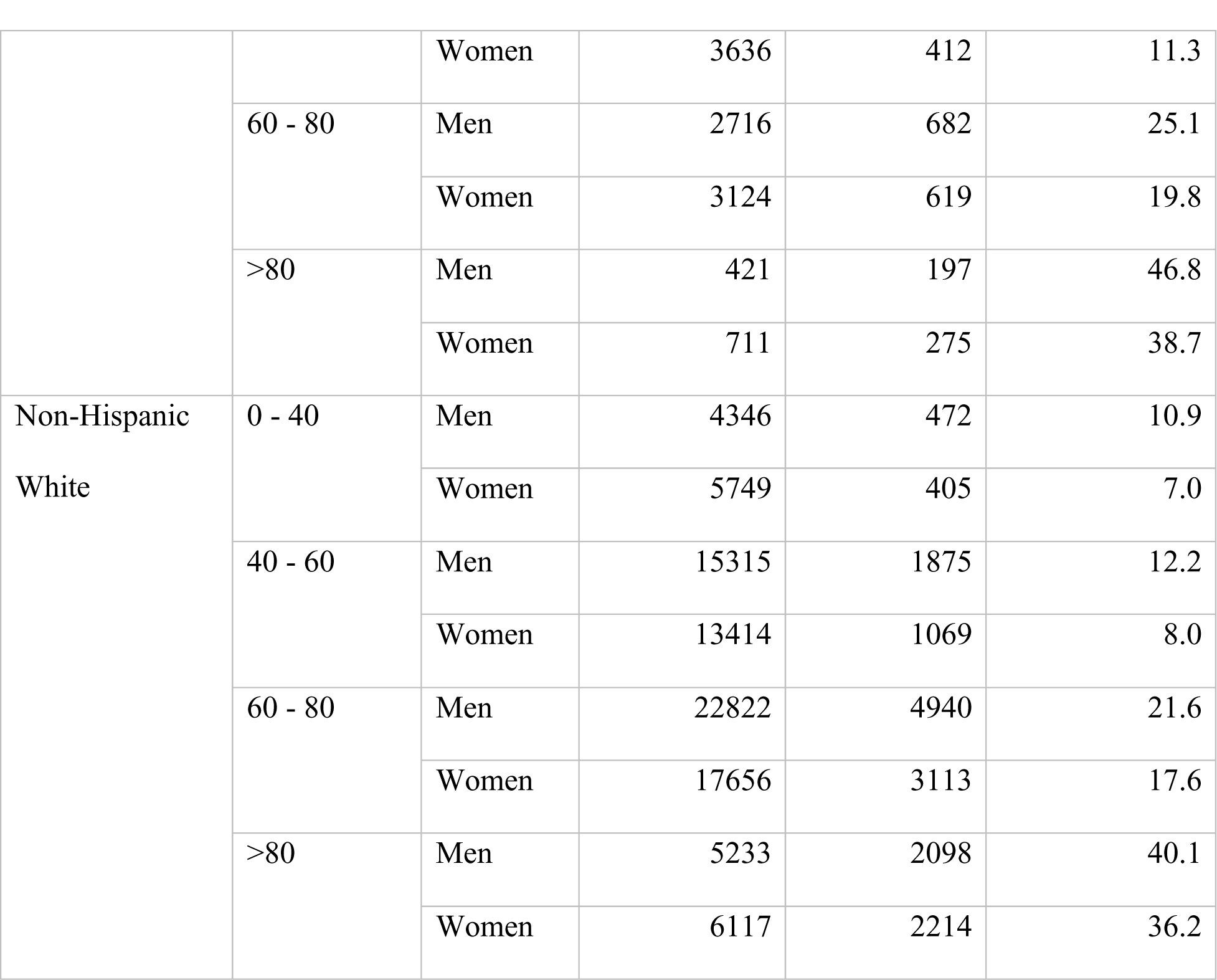
Demographics of heart failure cases by race, gender, and age

**Supplementary Table 2.**
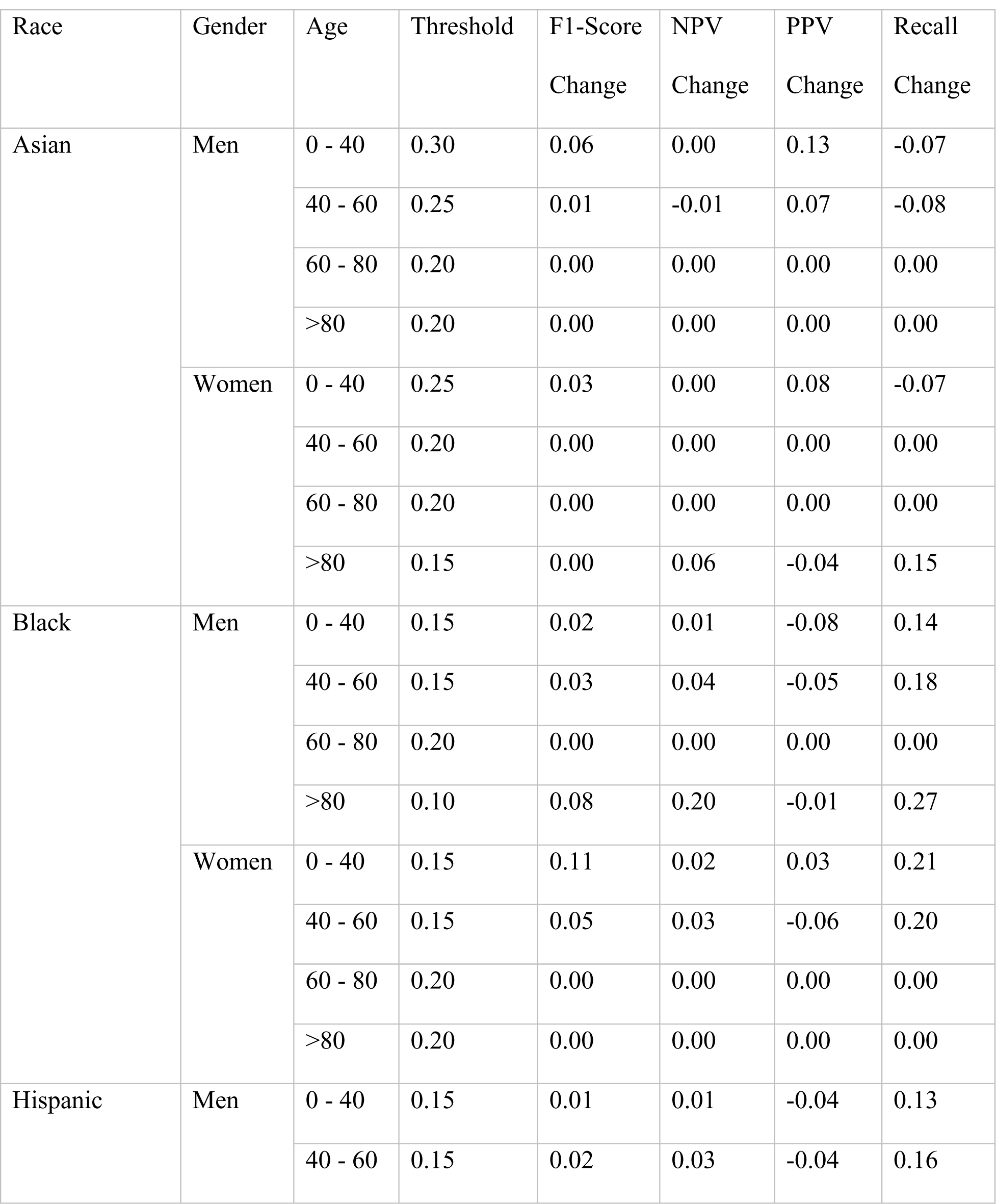
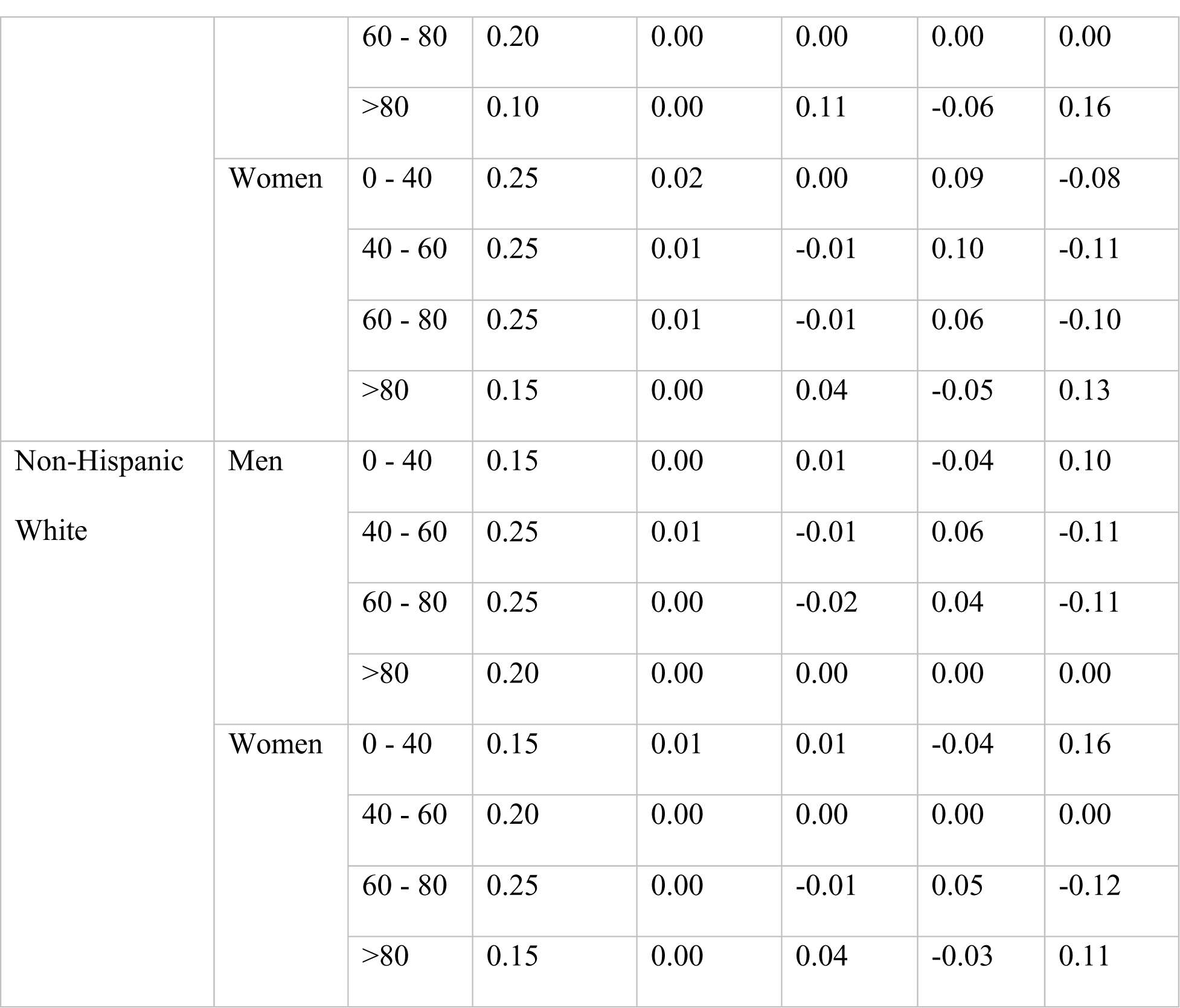
Changes in positive predictive value (PPV), negative predictive value (NPV), and recall using thresholds for binary cutoff optimized via individual F1-score vs overall optimal threshold (0.20)

**Supplementary Figure 1.**
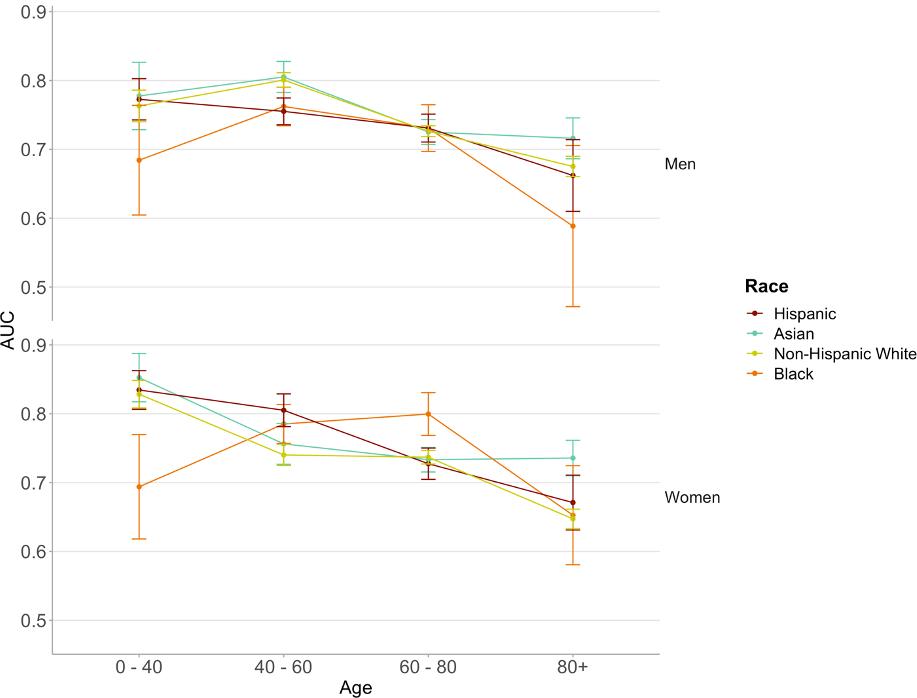
Age-related AUC trends stratified by race and gender

**Supplementary Figure 2.**
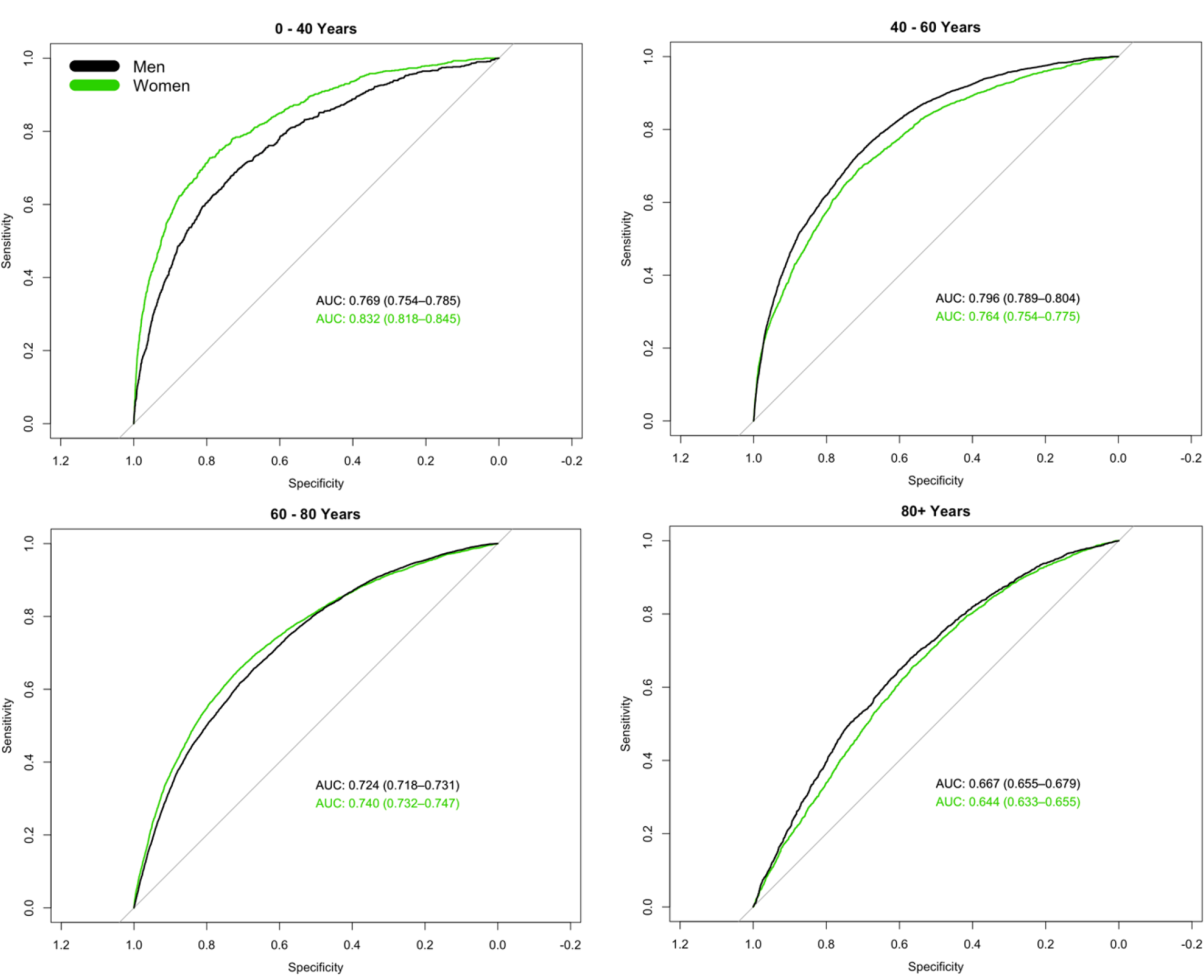
AUC curves stratified by gender across age groups

**Supplementary Figure 3.**
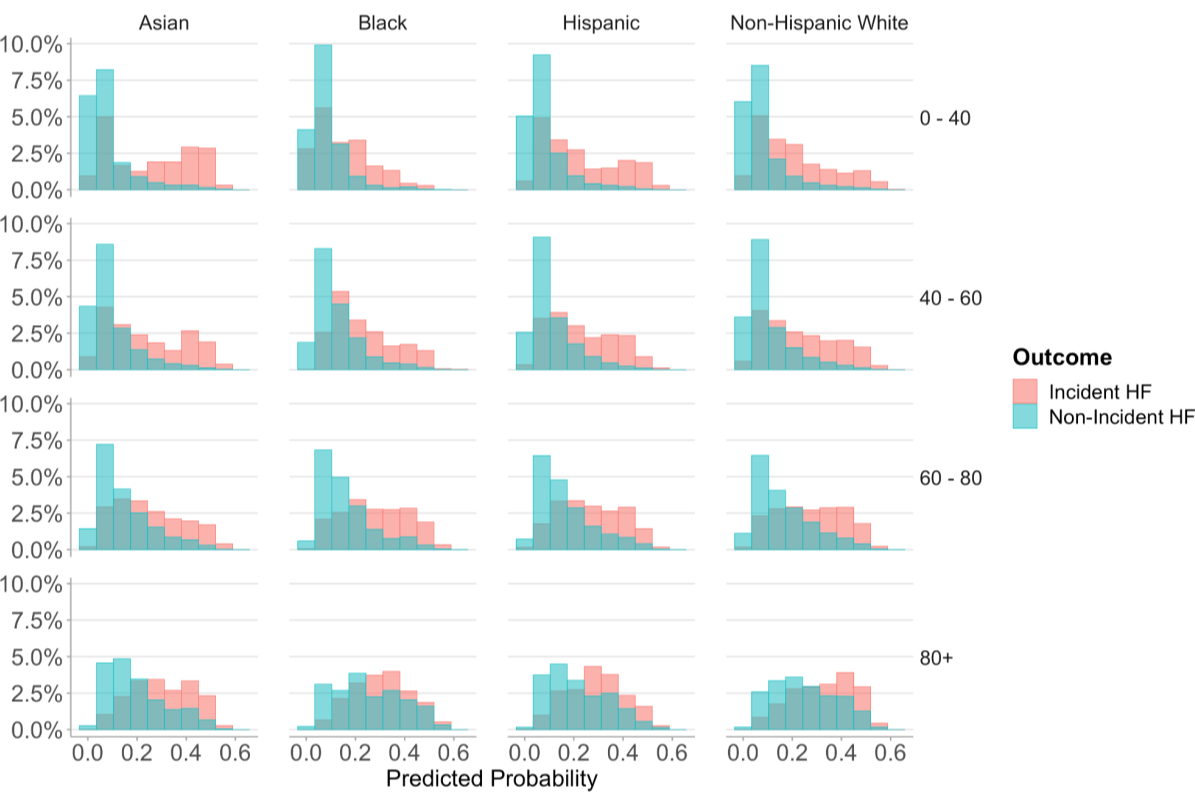
Probability distributions of cases with and without incident heart failure by race and gender

**Supplementary Figure 4.**
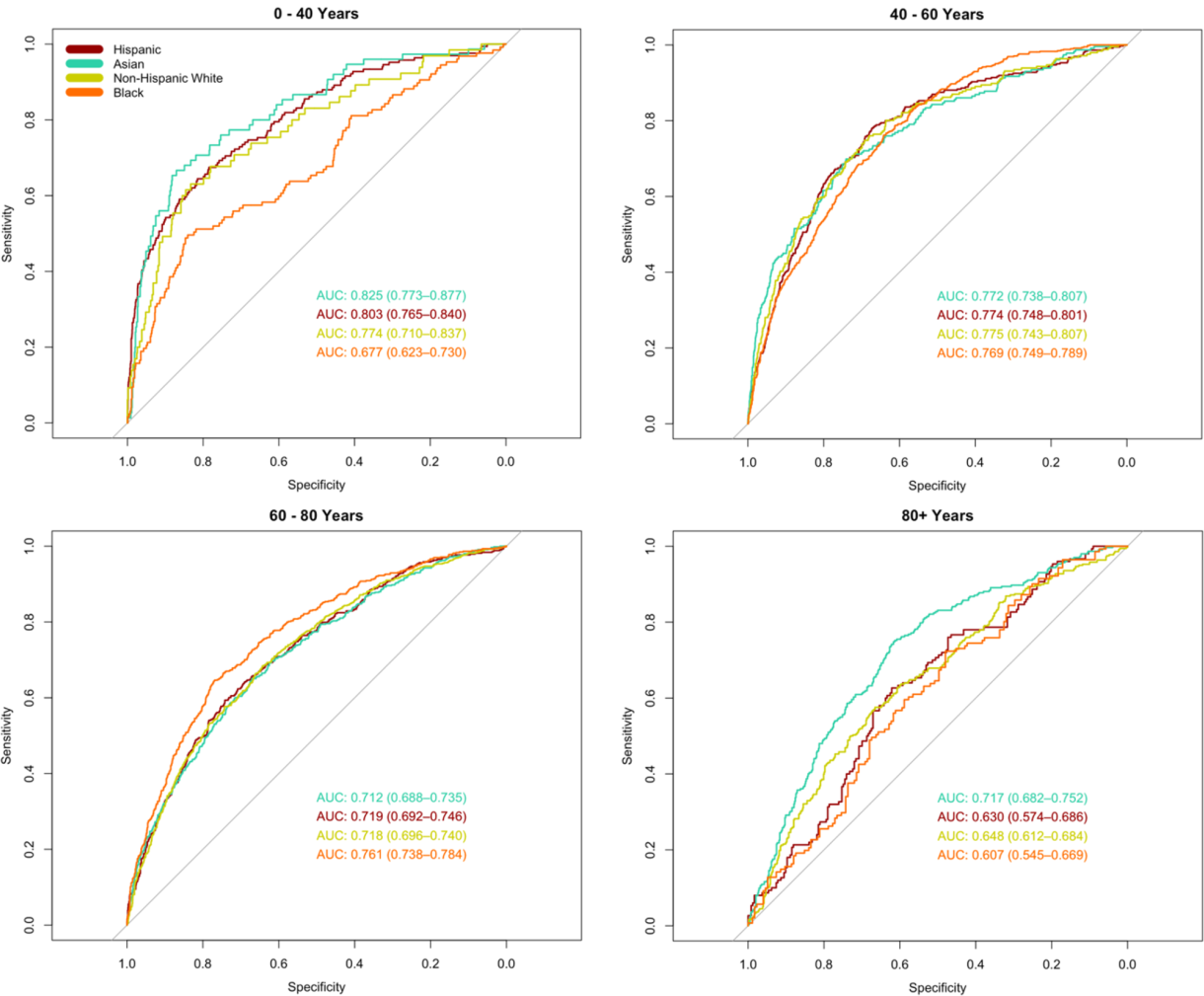
ROC curves stratified by race and age from a model trained on a dataset with equal racial representation.

**Supplementary Figure 5.**
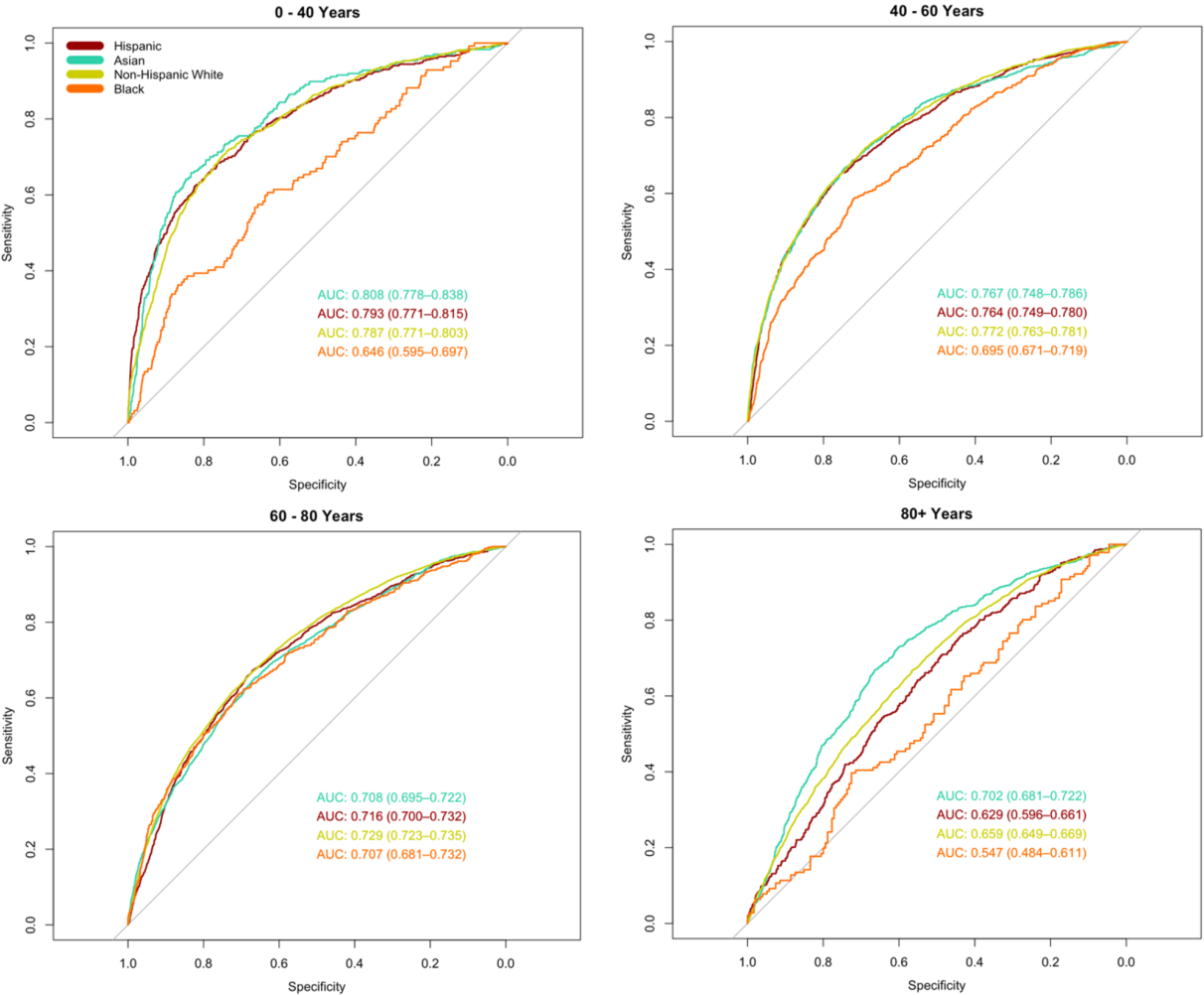
ROC curves stratified by race and age from models trained separately for each racial group.

**Supplementary Figure 6.**
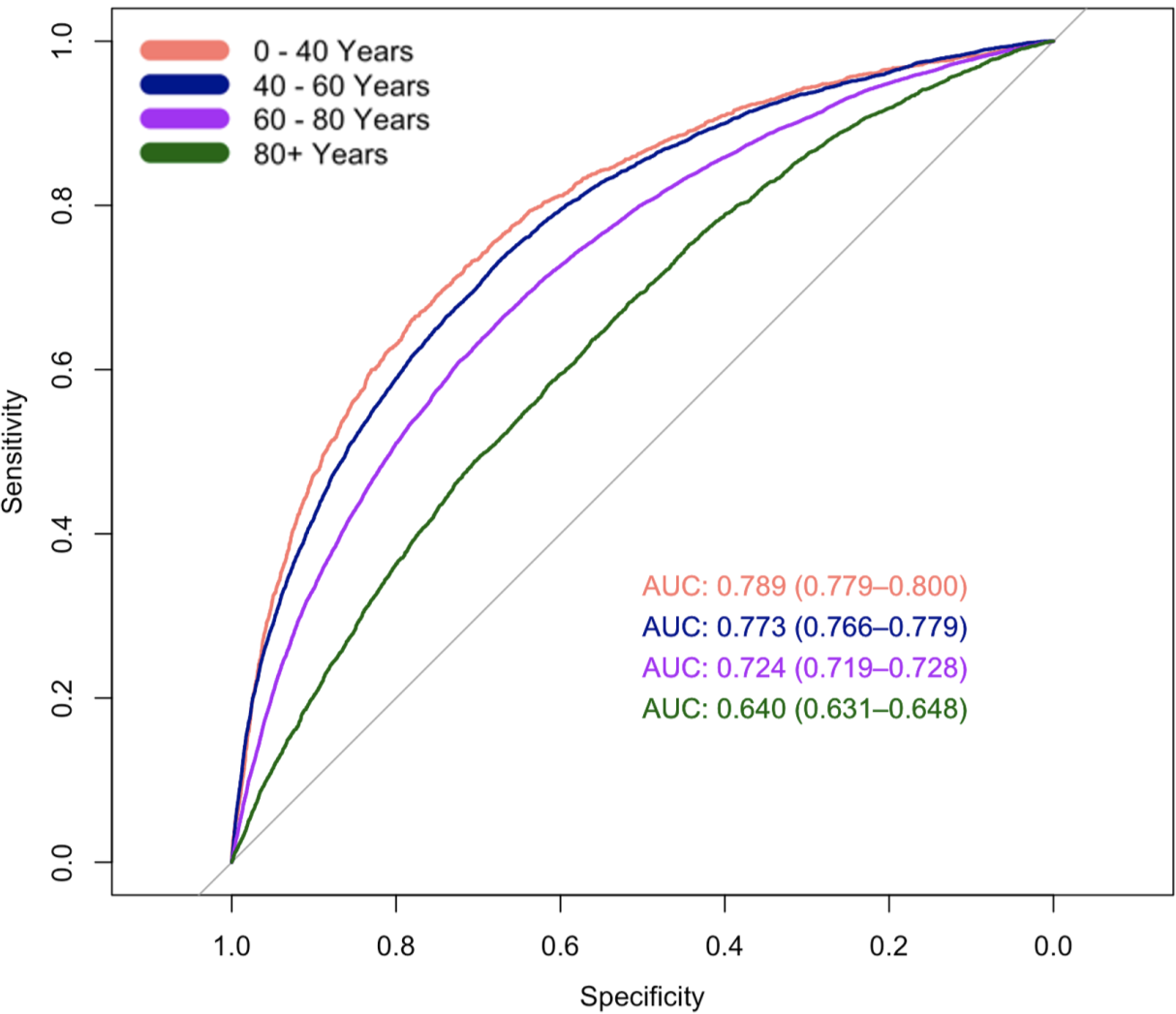
ROC curves from models trained and tested separately by age group

**Supplementary Figure 7.**
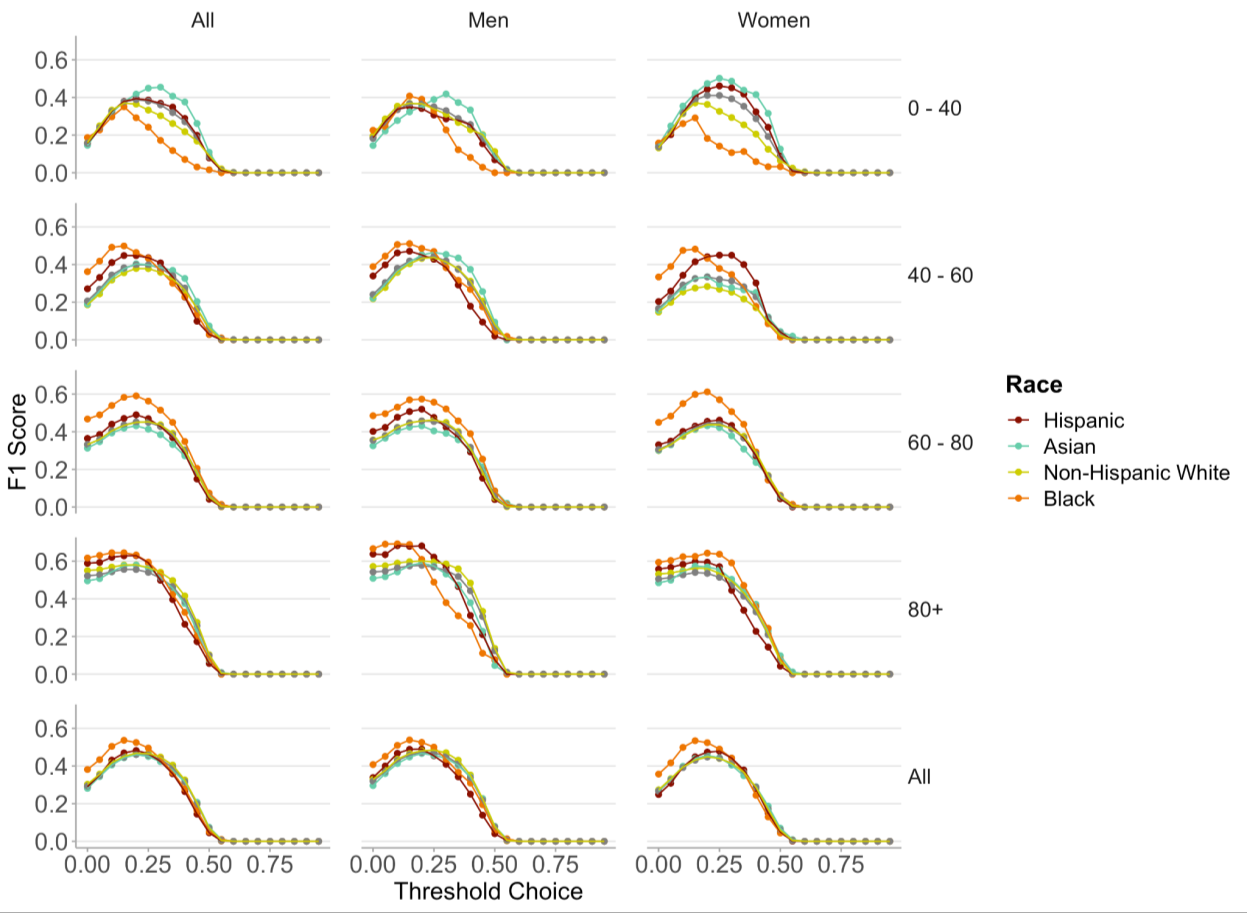
Optimizing threshold for F1-score by subgroups of race, age, and gender

